# CRISPR-based editing strategies to rectify *EYA1* complex genomic rearrangement linked to haploinsufficiency

**DOI:** 10.1101/2023.11.14.23298277

**Authors:** Yi Hwalin, Yejin Yun, Won Hoon Choi, Hye-Yeon Hwang, Ju Hyuen Cha, Heeyoung Seok, Jae-Jin Song, Jun Ho Lee, Seung Ha Oh, Sang-Yeon Lee, Daesik Kim

## Abstract

Pathogenic structure variations (SVs) and genomic rearrangements are associated with various types of cancer and rare genetic diseases. Recent studies have used Cas9 nuclease with paired guide RNAs (gRNAs) to generate targeted chromosomal rearrangements. Studies on Cas9-mediated translocations and inversions have mainly focused on producing fusion proteins that cause cancer, whereas research on precision genome editing for rectifying SVs is limited. In this study, through whole- genome sequencing, we identified a novel complex genomic rearrangement (CGR), specifically an *EYA1* inversion with a deletion, implicated in branchio-oto- renal/branchio-oto (BOR/BO) syndrome. The CGR results in a loss-of-function allele, leading to haploinsufficiency. To address this, two CRISPR-based editing approaches were tested. First, we engineered Cas9 nuclease and paired gRNAs tailored to the patient’s genome. The dual CRISPR/Cas9 system induced efficient editing at sites with paracentric inversion in patient-derived fibroblasts (up to 1.6%), and effectively restored the expression levels of the *EYA1* gene and its downstream targets, restoring overall transcriptional functionality. Additionally, we engineered gene-activating CRISPR-Cas modules (CRISPRa), which increased *EYA1* mRNA and protein expression to wild-type levels in human *EYA1* monoallelic knockout cells that mimic the haploinsufficiency. Moreover, CRISPRa significantly improved transcriptional activity essential for target gene expression. This suggests that CRISPRa-based gene therapies may offer substantial translational potential for approximately 70% of disease-causing *EYA1* variants responsible for haploinsufficiency. In parallel to deciphering the complexities of the genomic landscape related to human genetic disorders, our findings demonstrate the potential of CIRSPR-guided genome editing for correcting SVs, including those with *EYA1* CGR linked to haploinsufficiency.

## Introduction

Pathogenic structure variations (SVs) and genomic rearrangements are observed in various types of cancer and rare genetic diseases^1^. Genomic rearrangements are alterations in the architecture of genomic DNA that can result in complex structures such as inversions, deletions, duplications, and translocations^1^. These genomic rearrangements can arise from DNA double-strand breaks (DSBs) and incorrect rejoining of the DNA ends via mechanisms such as non-allelic homologous recombination and non-homologous end joining^1^. Chromosome microarrays and multiplex ligation-dependent probe amplification (MLPA) have become routine real- world techniques for detecting abnormal copy numbers^2^, but genomic approaches cannot identify balanced SVs, such as inversions and translocations. With the diagnostic and technical availability of whole-genome sequencing (WGS), a range of SVs, including complex genomic rearrangements (CGRs), can be identified at a much higher resolution than previously^3^, which leads to a better understanding of their mechanisms and potential therapeutic targets.

The CRISPR/Cas9 system has been adapted for site-specific genome editing in diverse cell types and model organisms^4,5^. Cas9 nuclease generates a DNA DSB at target sites, which induces the cellular repair process through either homology-directed repair or non-homologous end-joining (NHEJ) pathways. The development of advanced CRISPR nuclease using paired guide RNAs (gRNAs) achieves precise targeted gene deletions and replacement in human cells^6^. Furthermore, recent studies have used Cas9 nuclease in combination with paired gRNAs to generate targeted chromosomal rearrangements, focusing on cancer-associated chromosomal rearrangements such as *EML4-ALK, NPM-ALK, CD74- ROS1, KIF5B-RET, EWSR1-FLI1,* and *AML1-ETO*^7–10^. Previous studies have primarily focused on generating fusion proteins implicated in cancer, whereas the field of precise genome editing for correcting SVs requires further exploration.

Through WGS, we identified a novel CGR, characterized by an *EYA1* inversion with a deletion, responsible for branchio-oto-renal/branchio-oto (BOR/BO) syndrome. The CGR mutation results in a loss-of-function allele and thus haploinsufficiency served as the underlying mechanism. To rectify this, we developed two genome editing approaches. The Cas9 nuclease and paired gRNAs precisely induce the paracentric inversion to correct the CGR mutation with relatively high efficiency in patient-derived fibroblasts. This dual CRISPR/Cas9 system successfully restores the expression level of the *EYA1* gene, coupled with the expression of downstream target genes, leading to an improvement in transcriptional activity. In addition, through CRISPR activator (CRISPRa) with a versatile therapeutic profile, the expression level of both the *EYA1* gene and its encoded protein were increased similar to wild-type levels in human *EYA1* monoallelic knockout cells that mimic haploinsufficiency. The CRISPRa system significantly improved transcriptional activity essential for target gene expression, suggesting such advancements might be relevant to all disease-causing variants associated with *EYA1* haploinsufficiency.

## Materials and methods

### Participants

All procedures were approved by the Institutional Review Board of Seoul National University Hospital (IRB-H-0905-041-281 and IRB-H-2202-045-1298). In this study, one BOR/BO multiplex family segregated with CGR was included in the Hereditary Hearing Loss Clinic within the Otorhinolaryngology division of the Center for Rare Diseases, Seoul National University Hospital, Korea. The demographic data and clinical phenotypes were retrieved from electronic medical records. The presence and severity of associated medical conditions were determined using the Tenth Revision of the International Statistical Classification of Diseases and Related Health Problems (ICD-10) codes and/or features in their clinical manifestations.

### Whole-exome sequencing and Multiplex Ligation-dependent Probe Amplification

Genomic DNA was extracted from peripheral blood samples and used in whole- exome sequencing (WES) via SureSelectXT Human All Exon V5 (Agilent Technologies, Santa Clara, CA, USA). Adhering to the instructions provided, we prepared a library which was then sequenced using a NovaSeq 6000 system (Illumina, San Diego, CA, USA) in a paired-end manner. Sequence reads were compared to the human reference genome (GRCh38) and processed in line with the Genome Analysis Toolkit best-practice guidelines to identify single nucleotide variants (SNVs) and indels^11^. The ANNOVAR program was used for variant annotation, such as from the RefSeq gene set and gnomAD^12,13^. Rare non-silent variants were selected as candidates, including nonsynonymous SNVs, coding indels, and splicing variants. We also used the KRGDB and KOVA databases for further filtration of ethnic-specific variants^14,15^. We conducted a comprehensive bioinformatics analysis to detect candidate variants using a defined filtering process, as described previously^16–18^.

We also assessed the copy number status of *EYA1* using a SALSA Multiplex Ligation-dependent Probe Amplification (MLPA) P461 DIS Probemix kit (MRC- Holland, Amsterdam, Netherlands). The amplification products were analyzed with an ABI PRISM 3130 Genetic Analyzer (Applied Biosystems, Foster City, CA, USA), and the results were interpreted with the aid of Gene Marker 1.91 software (SoftGenetics, State College, PA, USA).

### Whole-genome sequencing and bioinformatics

Genomic DNA was extracted from peripheral blood samples using Allprep DNA/RNA kits (Qiagen, Venlo, Netherlands) and libraries were generated with TruSeq DNA PCR-Free Library Prep Kits (Illumina). The libraries were then sequenced on the Illumina NovaSeq6000 platform with the coverage set at an average depth of 30×. The obtained sequences were aligned to the human reference genome (GRCh38) using the BWA-MEM algorithm and PCR duplicates were eliminated using SAMBLASTER. Mutation calling for base substitutions and short indels was achieved with HaplotypeCaller2 and Strelka2, respectively. Delly was used to identify SVs. The breakpoints of the genomic rearrangements of interest were visually examined and validated. Variant filtering and assessment of their Mendelian inheritance patterns were carried out. The pathogenicity of the variants was classified using the American College of Medical Genetics and Genomics/Association for Molecular Pathology guidelines^19^.

### Cell culture and transfection

Patient fibroblasts were maintained in DMEM supplemented with 10% fetal bovine serum and 1% penicillin/streptomycin/amphotericin. To induce the correction of pathogenic inversion, 2×10^5^ patient fibroblasts were transfected with 17 μg Cas9 protein, 5 μg of in vitro transcribed gRNA targeting the A-B junction, and 5 μg of in vitro transcribed gRNA targeting the B-C junction using an Amaxa P3 Cell Line 4D- Nucleofector Kit (CM-137 program). Cells were analyzed 3 days after transfection.

HEK293T cells (American Type Culture Collection, ATCC, CRL-11268) were maintained in DMEM supplemented with 10% fetal bovine serum and 1% penicillin/streptomycin. For dCas9-VP64 mediated *EYA1* activation, HEK293T cells were seeded onto 24-well plates and transfected with 2,000 ng of plasmid DNA encoding dSaCas9-VP64 and gRNA (Addgene plasmid #158990) using 3 μL Lipofectamine 2000 (Life Technologies). After 72 h, total RNA was isolated with an RNeasy Mini Kit (Qiagen) according to the manufacturer’s instructions.

### Targeted deep sequencing

Genomic DNA containing the on-target was amplified using KAPA HiFi HotStart DNA polymerase. The amplified products were designed to include Illumina TruSeq HT dual index adapter sequences. Subsequently, the amplified products were subjected to 150-bp paired-end sequencing using the Illumina iSeq 100 platform. To calculate the frequencies of insertions and deletions (indels), we used the MAUND tool, which is available at https://github.com/ibs-cge/maund.

### Digital droplet PCR

Molecular analysis of the CGR derived from WGS was conducted using breakpoint PCR and digital droplet PCR (ddPCR). Genomic DNA extracted from patients was used to perform ddPCR with the QX200 ddPCR system (Bio-Rad). Procedures followed the manufacturer’s protocol with minor modifications. Briefly, the master mix for ddPCR consisted of 100 ng of genomic DNA, 1×ddPCR supermix for probes (no dUTP; 186-3023, Bio-Rad), 1.0CμM primer, and 0.25CμM probe (Metabion, Germany). Primers and a probe were designed for *EYA1* breakpoint junctions **(Supplementary Table 1)**. The samples were thoroughly combined before being put into a Bio-Rad QX100/QX200 droplet generator’s DG8 cartridge. The cartridge was then put into the QX200 Droplet Generator (Bio-Rad) and Droplet Generation Oil was added. After droplet formation, the droplets were carefully moved to a twin-tec semi-skirted 96-well PCR plate (EP0030128575, Eppendorf). PCR was then carried out in a C1000 Touch thermal cycler (Bio-Rad) for the two-step running program of 94°C for 30s and 56 °C for 1min for 40 cycles with an initial and final incubation at 95°C and 98 °C, respectively. The signal in each droplet was read by using a droplet reader and analyzed using the QuantaSoft program (Version 1.7, Bio-Rad).

### RNA isolation and real-time quantitative PCR

Total RNA (1 μg) was extracted from either fibroblasts or HEK293T cells using the TRIzol method, with subsequent purification via Rneasy mini-columns (Qiagen) and an incorporated on-column Dnase I treatment. The synthesis of cDNA was achieved from 2 μg of the isolated total RNA using the reverse transcription (RT)-PCR method and Accupower RT-pre-mix (Bioneer). Quantitative RT-PCR assays were performed on cDNA samples diluted 1/20 using SYBR qPCR master mix (Kapa Biosystems, USA) as the reporter dye. Primers at a concentration of 10 pmol were used to detect specific gene mRNA expression. Their sequences were as follows: *SOX2*, forward 5′-GCTACAGCATGATGCAGGACCA-3′ and reverse 5′- TCTGCGAGCTGGTCATGGAGTT-3′; *NEUROG1*, forward 5′-GCCTCCGAAGACTTCACCTACC-3′ and reverse 5′- GGAAAGTAACAGTGTCTACAAAGG-3′; *GFI1*, forward 5′-GCTTCAAGAGGTCATCCACACTG3′ and reverse ACCTGGCACTTGTGAGGCTTCT-3′; and *GAPDH*, forward 5′- GAGTCAACGGATTTGGTCGT-3′ and reverse 5′- GACAAGCTTCCCGTTCTCAG-3′.

The primer sequences for *EYA1* can be found in the Supplementary tables. The relative frequency of *EYA1* mRNA was determined using the comparative C_T_ method^20^.

### Luciferase reporter gene assay

The luciferase reporter gene assay was performed, as described previously with slight modifications^17^. Briefly, HEK293T cells were transfected initially with three distinct plasmids: pGL4.12[luc2CP]-MYOG-6xMEF3, pRK5-SIX1, and pRL/CMV (E2261, Promega). Post-transfection, the cells were harvested for a luciferase assay using a Dual-Luciferase Reporter Assay kit (E1910, Promega), following the manufacturer’s guidelines. Transfection efficiency was adjusted based on Renilla activity, which was assessed after co-transfection with pRL/CMV.

### Generating single cell-derived *EYA1* knockout clones

HEK293T cells were maintained in DMEM supplemented with 10% FBS and 1% penicillin-streptomycin. For the transfection, the cells were seeded in 24-well plates and co-transfected with 1500 ng of the Cas9 expression plasmid and 500 ng of a sgRNA-encoding plasmid containing a spacer sequence (5′- ATGCCACGTACCCACAGCC-3′ on the top strand and 5′- GGCTGTGGGTACGTGGCAT-3′ on the bottom strand). Single-cell clones were generated via limiting dilutions in 96-well plates, followed by clonal expansion. Genomic DNA from these clones was isolated using Allprep DNA/RNA kits (Qiagen) according to the manufacturer’s instructions, and mutation frequencies were determined through targeted deep sequencing.

### Immunoblotting

Cells were washed with PBS and lysed using RIPA buffer supplemented with protease inhibitors. The lysates combined with sample buffer were denatured at 85°C in preparation for SDS-PAGE. Subsequently, proteins were transferred to polyvinylidene difluoride (PVDF) membranes. Membranes were blocked in 5% nonfat milk in TBS-T and then probed with primary antibodies. After comprehensive washing, membranes were exposed to horseradish peroxidase-linked secondary antibodies, specific to rabbit or mouse IgG. Protein bands became evident upon chemiluminescence application and were subsequently imaged. Band intensities were quantified using ImageJ software based on replicated experiments. Statistical evaluation was conducted using GraphPad Prism V5. Results with a p-value < 0.05 were used to confirm significance. Sources of antibodies and working dilutions were as follows: beta-actin, anti-β-actin (A1978, Sigma, 1/10,000); anti-EYA1 (22658-1-AP, Proteintech, 1/1,000).

## Results

### Identification of a novel complex genomic rearrangement in the *EYA1* gene

The proband, mother of the proband and affected siblings in SNUH family exhibited a range of clinical phenotypes associated with BOR/BO syndrome, demonstrating complete penetrance but variable expressivity of clinical phenotypes (**Figure 1a**). A bioinformatics approach and filtering strategy of WES results, aimed at prioritizing candidate causal variants for BOR/BO syndrome, yielded negative results. The sequential MLPA test showed a normal copy number state in *EYA1* (**Supplementary Figure 1**). A thorough examination of a region on chromosome 8 was conducted by leveraging SV profiles via family-based WGS. This process revealed a novel CGR involving an *EYA1* inversion (g.71211857 to g.71228236; with a 5′-breakpoint in intron 16 of *EYA1* and a 3′-breakpoint in intron 12 of *EYA1*), with a deletion at the end of the segment (g.71211857 to g.71215145; with both the 5′-breakpoint and 3’- breakpoint located in intron 16) (**Figure 1b and Supplementary Figure 2**). We identified two neo-junctional reads, one with 1 bp of microhomology (A-B, g.71215145) and the other with 2 bp of microhomology (B-C, g.71228236) (**Supplementary Figure 3**). To confirm the breakpoint junctions identified by WGS, we designed four pairs of forward and reverse primers (**Supplementary Table 1**). Gap-PCR revealed the amplification of a 954-bp PCR product corresponding to the breakpoint within intron 16 (A-B junction) and a 235-bp PCR product corresponding to the breakpoint within intron 12 (B-C junction) (**Figure 1c**). We also verified the genomic sequence of junctions by Sanger sequencing (**Figure 1c**). The CGR in *EYA1* results in a frameshift variant, leading to a premature stop codon at codon CTA within exon 16. This stop codon results in a truncated non-functional protein that lacks 210 amino acids at the C-terminal end, a region that includes the functional EYA domain (**Figure 1d**).

**Figure 1.**
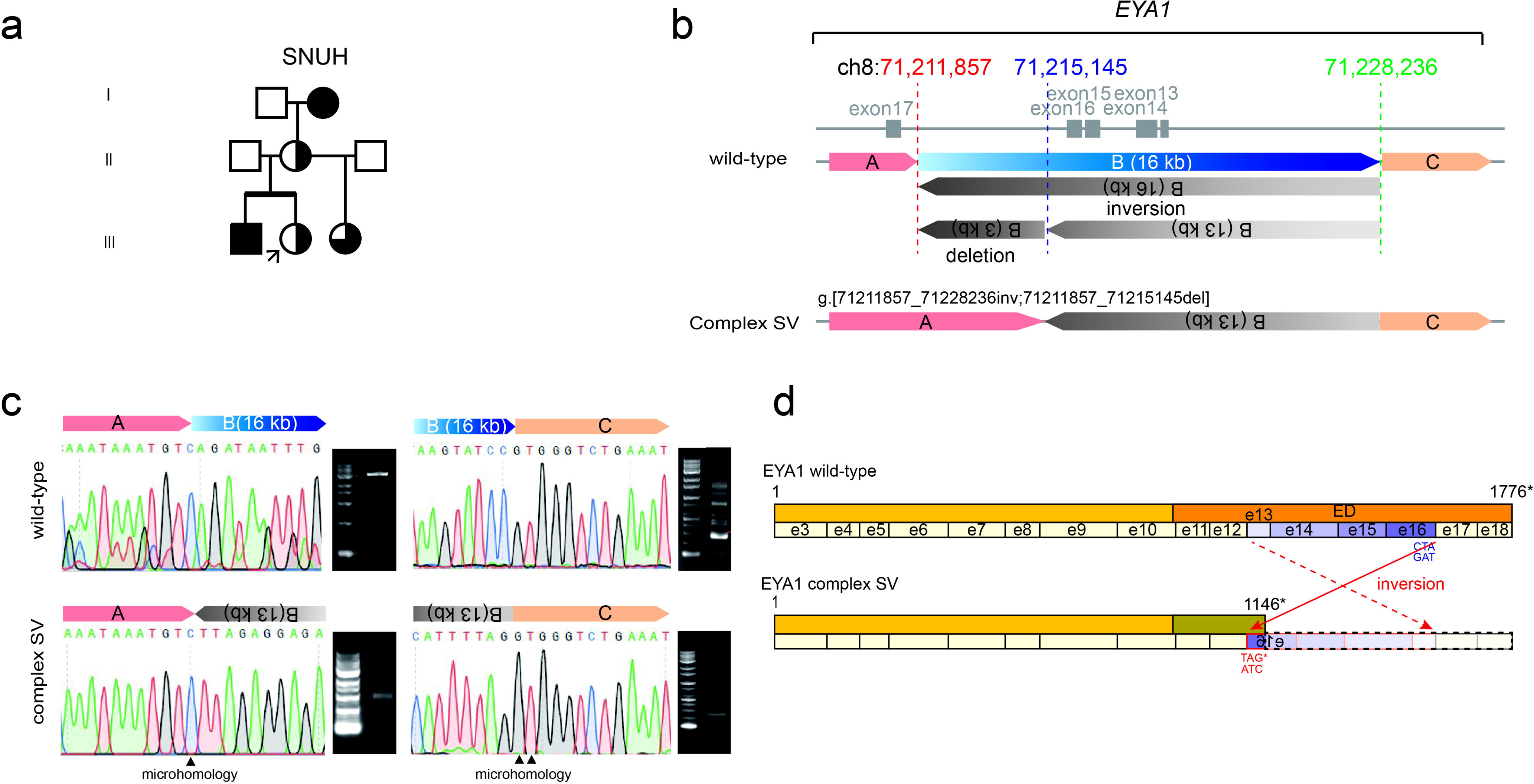
Pedigrees, clinical phenotypes, and two novel *EYA1* SVs harboring BOR/BO syndrome. (a) Pedigree of the SNUH family with BOR/BO syndrome. The symbols in the pedigree indicate the following: circle, female; square, male. (b) Schematic representation illustrating the orientation and location of genomic segments reconstructed with the complex genomic rearrangement (CGR). The genomic coordinates of the breakpoints (depicted by red, blue, and green dotted vertical lines) demarcate the junctions of the segments. The colors of the bars on the breakpoint junction correspond to the following genomic positions: red, 71197433-71211857; blue, 71211857-71228236; orange, 71228236-71548094. The coding sequence configuration is depicted as connected segments. (c) Gap-PCR results align with the breakpoints identified through WGS. Sanger sequencing verified the generated PCR products. (d) The predicted effect of the *EYA1* CGR on protein and domain structure. The solid red arrow represents the position of the inverted exon 16 (e16), whereas the dotted arrow denotes the position of the inverted exon 13 (e13). The abbreviations used in the domain map are defined as follows: e, exon; ED, EYA domain

### Cas9 nuclease with paired gRNAs induce efficient editing at sites of the paracentric inversion

To assess the potential of Cas9 nucleases with paired gRNA for correcting the pathogenic inversion in patient-derived fibroblasts (**Figures 2a and 2b**), we designed gRNAs that specifically target the junction sites between fragment A and the inverted B (13 kb) fragment, as well as between the inverted B (13 kb) and C fragments (**Figure 2c**). We transfected patient fibroblast cells with complexes of Cas9 protein and *in vitro* transcribed gRNAs targeting the fragment junctions (**Figure 2c**), and measured editing frequencies using targeted deep sequencing (**Figure 2d**). The editing efficiency at target sites AB-T1 and AB-T2, located at the junction between fragment A and the inverted B (13 kb) fragment (A-B junction), were found to be 36.9% and 1.9%, respectively. Similarly, the Cas9 nuclease targeting the junctions between the inverted B (13 kb) fragment and fragment C (B-C junction) at sites BC-T1, BC-T2, and BC-T3 showed editing efficiencies of 57.2%, 89.0%, and 78.6%, respectively. Based on these results, we selected Cas9 nucleases targeting either AB-T1 and BC- T2 or AB-T1 and BC-T3 to induce the correction of the pathogenic inversion.

**Figure 2.**
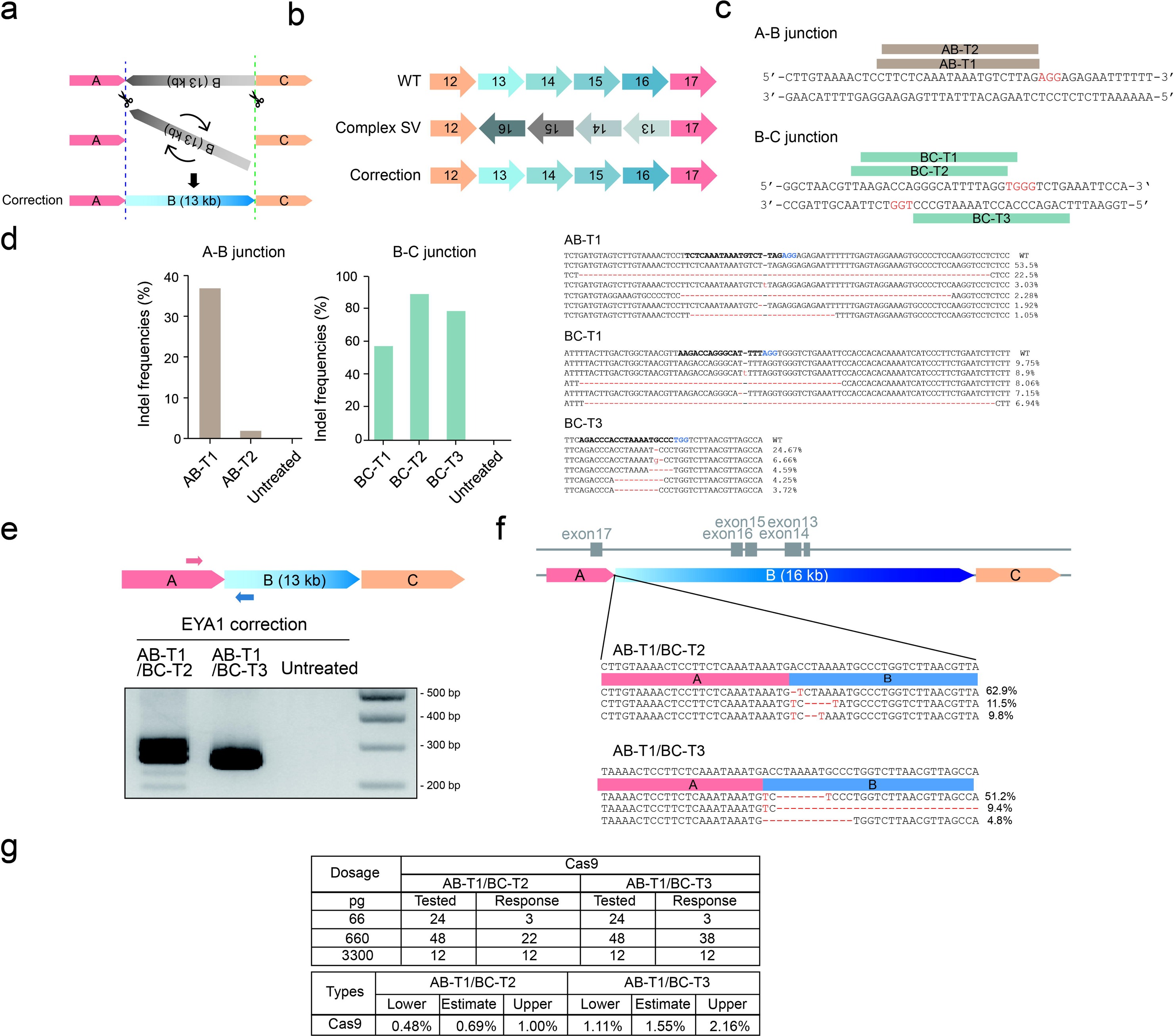
Dual Cas9 nuclease mediated correction of *EYA1* pathogenic complex. (a) Schematic representation of Cas9 nuclease and paired gRNA-mediated correction of the pathogenic inversion. The dashed lines represent the Cas9 target sites. (b) Schematic representation of the mature mRNA anticipated after correction of the pathogenic inversion. (c) The Cas9 target sequences at the A-B junction and B-C junction of the pathogenic *EYA1* gene in patient fibroblasts. The protospacer adjacent motif is highlighted in red. (d) Mutation frequencies induced by Cas9 nuclease at the A-B junction and B-C junction of the *EYA1* gene. (e) Detection of Cas9 nuclease-mediated inversions using inversion-specific primers. (f) Targeted deep sequencing reads of amplicons generated using inversion-specific primers. (g) Estimation of inversion frequencies by digital PCR analysis using serially diluted samples. Diluted genomic DNA samples were subjected to PCR using inversion- specific primers. The results were analyzed using the Extreme Limiting Dilution Analysis program (http://bioinf.wehi.edu.au/software/elda/).

We next investigated whether Cas9 nuclease with paired gRNAs could correct the pathogenic inversion between endogenous loci in patient-derived fibroblasts (**Figure 2a**). To induce a paracentric inversion between the A-B junction and B-C junction, we co-transfected Cas9 protein and paired gRNAs targeting the two junctions. We then amplified the expected inversion junctions with paracentric inversion-specific primer pairs and found that inversions were induced in cells transfected with Cas9 protein and paired gRNAs **(Figure 2e)**. To further analyze these amplicons, we performed targeted deep sequencing and found that Cas9 nuclease could induce a paracentric inversion that corrects the pathogenic inversion with additional small insertions or deletions (indels) at the junctions in patient-derived fibroblast cells (**Figure 2f**). To measure the inversion frequencies induced by Cas9 nuclease and paired gRNAs, we used dilution PCR^21,22^. Through this analysis, we observed that the inversion frequency induced by Cas9 nuclease and paired gRNAs was up to 1.6% in patient-derived fibroblast cells (**Figure 2g**). These results demonstrate that Cas9 nuclease and paired gRNAs, which are specific to the patient’s genome, can correct the pathogenic inversions with relatively high efficiency in patient-derived fibroblasts.

### Cas9 nuclease with paired gRNAs restores *EYA1* expression and transcriptional activity

We assessed whether the Cas9 nuclease with paired gRNAs could restore the variant allele frequency (VAF) of the inversion junctions in patient cells using ddPCR (**Figure 3a**). We designed a primer/probe set that targeted both reference and mutated gene sequences at the A-B and B-C breakpoint junctions (**Figure 3b**). The VAF of the A-B and B-C junctions compared to the reference gene in patient fibroblast cells were 0.507 and 0.488, respectively. Notably, in the edited cells in which the AB-T1 and BC-T2 sites were targeted, the VAF of the A-B and B-C junctions decreased to 0.381 and 0.181, respectively. Similarly, the ratios dropped to 0.392 and 0.233 when AB-T1 and BC-T3 were targeted (**Figure 3c**). To verify that the genome editing elicited a recovery of the *EYA1* transcript, we conducted quantitative reverse transcription PCR (qRT-PCR) with a primer set that covered the breakpoint junctions (**Supplementary Table 3**). Targeting both combinations of sites (AB-T1 with BC-T2 and AB-T1 with BC-T3) led to a significant increase in the amount of wild-type *EYA1* transcripts (**fig. 3d**). To assess the potential restoration of the *EYA1* transcriptional activity, we used a luciferase assay (**Figure 3e**). EYA1 encodes a transcriptional coactivator; the Eya domain is crucial for the formation of the EYA1-SIX1-DNA complex that regulates the transcription of target genes involved in the development of the branchial arch, otic system, and renal system^23^. In this context, we used the pGL4.12[luc2CP]-MYOG-6xMEF3 construct, which incorporates a luciferase reporter along with six repeats of the MEF3 motif. Importantly, each motif exhibits a specific affinity for the EYA1 and SIX1 protein complex. Under the conditions of SIX1 overexpression, we observed that the luciferase activity in the *EYA1* mutant significantly decreased to 12.7% of the wild- type levels, representing a 7.8-fold reduction. On the other hand, cells with edited genes displayed a substantial increase in luciferase activity relative to the mutant, with increases of 2.7-fold at the AB-T1 and BC-T2 sites, and 3.4-fold at the AB-T1 and BC-T3 sites. These levels correspond to 34.1% and 43.8%, respectively, of those observed in control cells. Although it is essential to investigate the downstream target gene expression and the modifications in the transcriptome tied to *EYA1* transcriptional activity, our data suggest that the dual Cas9 nucleases, designed to specifically target the paracentric inversion in the patient genome, are capable of restoring the transcriptional functionality of *EYA1*.

**Figure 3.**
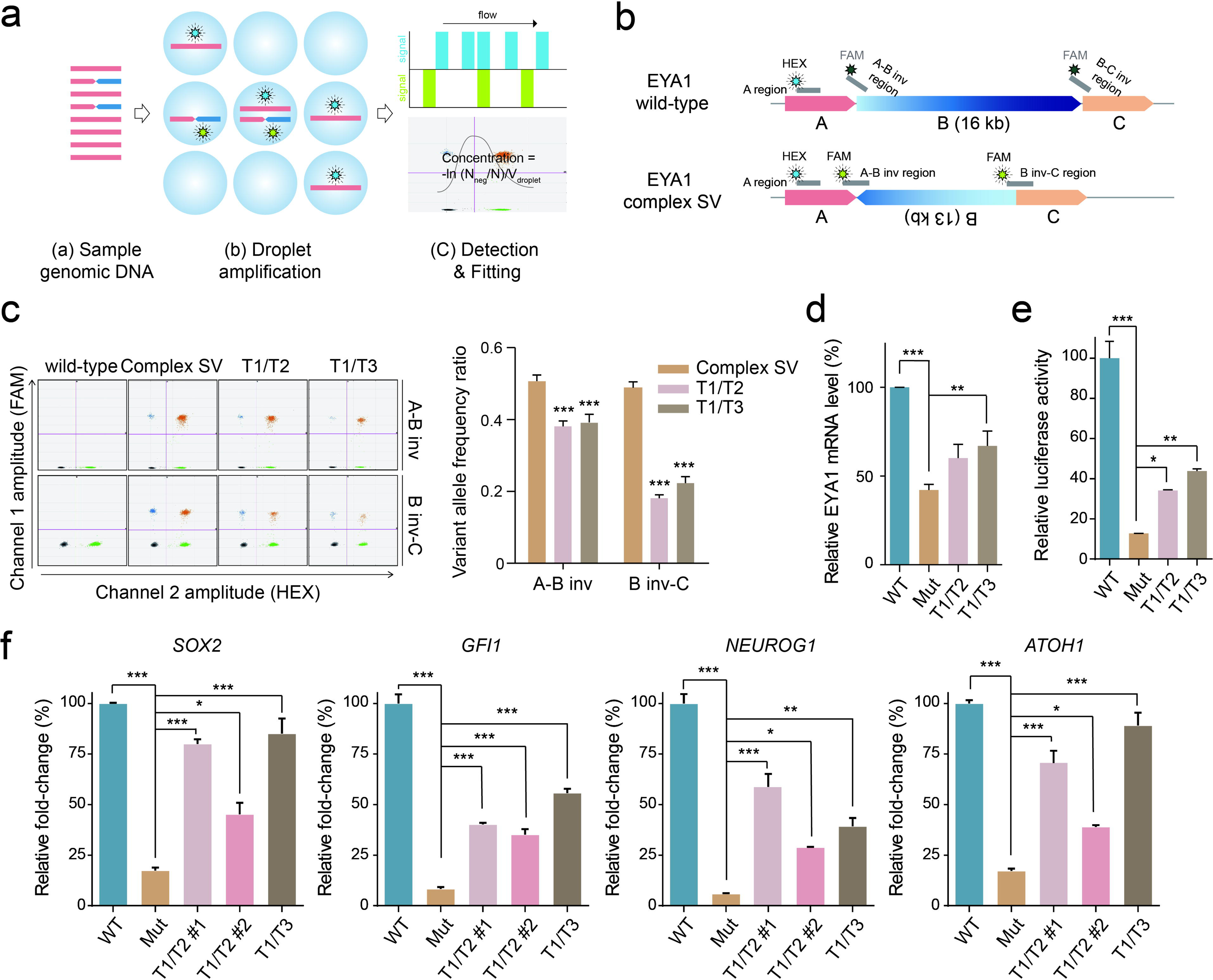
**Restoration of *EYA1* gene expression**(a) Schematic workflow of digital droplet PCR (ddPCR). The genomic DNA was subjected to amplification after partitioning into each droplet, see (a) and (b). Multiple fluorescent signals were detected to fit into the Poisson distribution, see (c), to calculate absolute concentration. (b) Primer and probe design. *EYA1* wild-type and complex SV-specific forward and reverse primers were used to verify the breakpoint junction. (c) Two-dimensional plots of ddPCR results represent the *EYA1* variant allele frequency at each breakpoint junction. The colored dots in the 2D plot indicate the following: gray, negative droplets; blue, droplets with mutant allele; green, droplets with wild-type allele; orange, droplets with both mutant and wild-type alleles. Genomic DNA was isolated from the *EYA1* wild-type (control), *EYA1* mutated (patient), and *EYA1*-edited fibroblast cells. *p* < 0.01*** (n = 3, one-way analysis of variance (ANOVA) followed by Bonferroni’s multiple comparison test). (d) Wild-type *EYA1* transcript levels in both patient-derived and edited fibroblast cells. Quantitative reverse transcription-polymerase chain reaction (qRT-PCR) analysis was performed, and the *GAPDH* mRNA levels were used to normalize the levels of *EYA1* expression. Each bar represents the means of percent values (relative to the *EYA1* mRNA levels in wild-type fibroblast cells) ± SEM from three independent experiments. (*, *p* < 0.05; **, *p* < 0.01 one-way ANOVA with Bonferroni *post hoc* test) (e) Fibroblast cells were concurrently transfected with an *MYOG* promoter-driven luciferase reporter and the *SIX1* wild-type construct. Relative luciferase activity (wild-type as control) was plotted as mean ± SEM (n = 3), *, *p* < 0.05, **, *p* < 0.01***, *p* < 0.001 (one-way ANOVA followed by Bonferroni’s multiple comparison test). (f) Total RNA was isolated, and the target genes (*SOX2*, *GFI1*, *NEUROG1*, and *ATOH1*) of *EYA1* were analyzed by qRT-PCR. *GAPDH* mRNA levels were used to normalize the results. Values are represented as the mean ± SEM of three independent experiments (N = 3); *, *p* < 0.05, **, *p* < 0.01, ***, *p* < 0.001 (n = 3, one-way ANOVA with Bonferroni’s multiple comparison test).

In the literature, *EYA1* CGRs involving inversions and deletions have been well-documented in BOR/BO cases^24,25^. Consistent with this, our in-house database revealed that two unrelated BOR/BO families were segregated with CGRs (**Supplementary Figure 2**), including a cryptic large inversion, accounting for 18% (**Supplementary Table 4**). Thus, the dual CRISPR/Cas9 system developed herein has the potential to correct pathogenic SVs, including those with *EYA1* CGRs, implicated in human genetic disorders.

### Development of *EYA1* CRISPR activator for haploinsufficiency

The pathogenic inversion occurring in the *EYA1* leads to nonsense-mediated mRNA decay, resulting in haploinsufficiency (**Figures 1d and 3d**). Notably, when the cells were treated with cycloheximide (CHX), stabilization of the *EYA1* transcript was observed, suggesting that the haploinsufficiency of *EYA1* is dependent on NMD (**Figure 4a**). To investigate the potential of CRISPRa for inducing increased endogenous *EYA1* expression, we transfected plasmid DNA encoding dSaCas9- VP64 and gRNA targeting the *EYA1* promoter into HEK293T cells and compared the relative expression of *EYA1* mRNA in treated cells and control cells (**Figure 4b**). Using qRT-PCR, we observed that dSaCas9-VP64 led to a substantial 4.0-fold upregulation of *EYA1* mRNA expression compared to untreated HEK293T cells. To model the potential haploinsufficiency originating from a variety of mechanisms and to evaluate the performance of the CRISPRa, we engineered HEK293T cells with either monoallelic or biallelic knockouts of *EYA1* (**Figure 4c**). We then investigated whether co-expressing dSaCas9-VP64 with either gRNA2 or 3 affected the mRNA expression of *EYA1* in these cell lines. Quantitative assessments of relative mRNA abundances indicated marked increases in wild-type and monoallelic knockout cells, with up to 3.7 and 1.4-fold increases, respectively, when compared to untreated cells. In contrast, cells with biallelic knockouts exhibited no discernible alterations in mRNA levels (**Figure 4d**). We further analyzed transcriptional activity of *EYA1* through a luciferase-based reporter gene assay, aiming to assess the potential restoration of *EYA1* functionality with CRISPRa (**Figure 4e**). Under normal conditions, the luciferase activity in both monoallelic and biallelic knockout cells showed substantial reductions, by 68.5% and 64.4%, respectively, compared to HEK293T cells. Conversely, cells subjected to CRISPRa had enhanced luminescence, with increases of 1.3-fold in HEK293 cells and 1.7-fold in monoallelic knockout cells relative to their untreated counterparts. Consistent with the results of mRNA levels, biallelic knockout cells were not affected by CRISPRa. In line with these observations, protein levels in CRISPRa-treated cells increased by 1.84-fold in HEK293T cells and 8.00-fold in monoallelic knockout cells (**Figure 4f)**.

**Figure 4.**
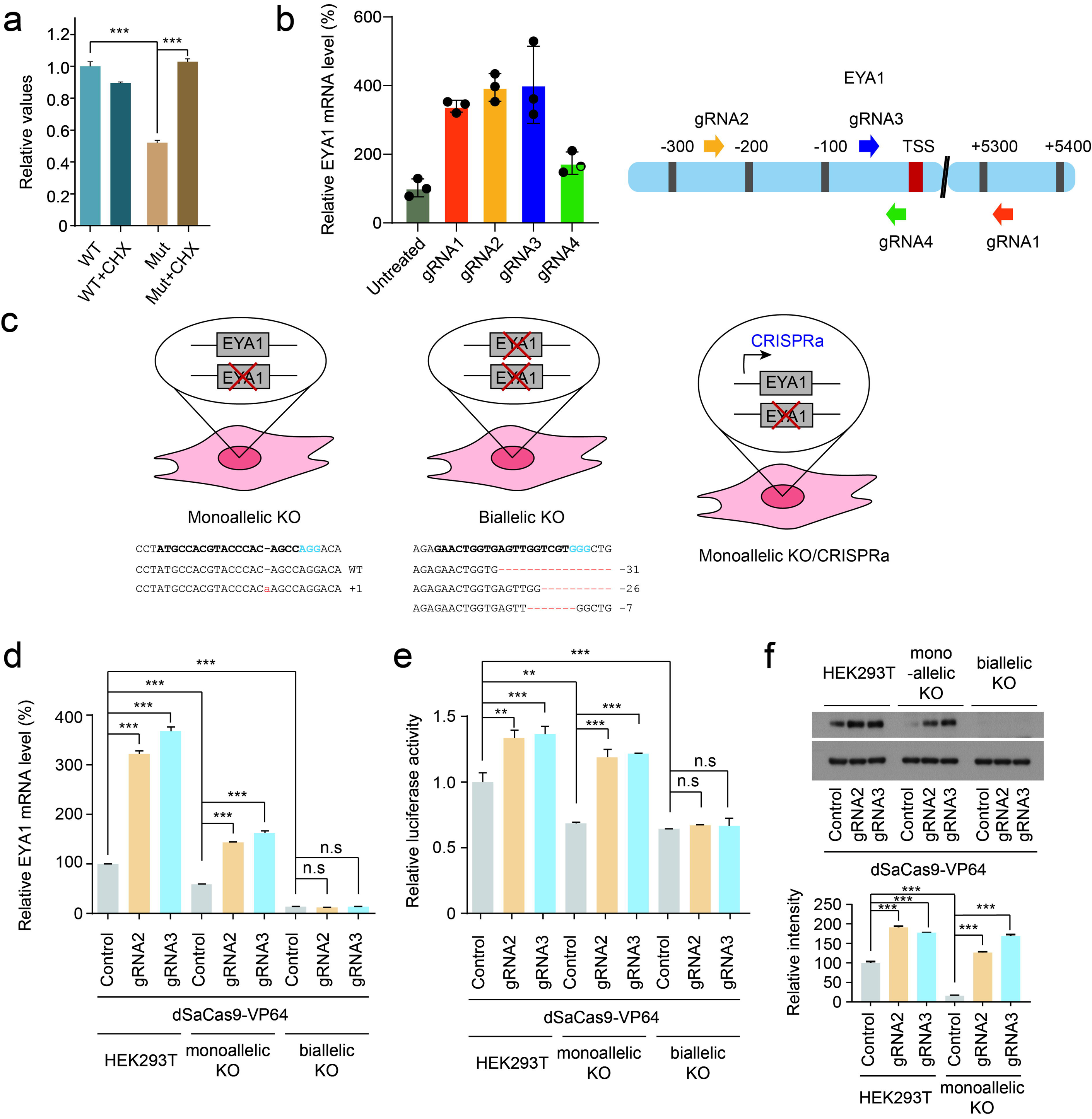
Designing a CRISPR activator strategy to elevate *EYA1* levels. (a) Relative *EYA1* transcript levels were determined by qRT-PCR. Fibroblast cells were treated with DMSO or CHX (50 μg/mL) for 6h. All values were normalized to the untreated wild-type cells data and plotted as mean ± SEM (n = 3), ***, *p* < 0.001 (one-way ANOVA followed by Bonferroni’s multiple comparison test). (b) (*left*) HEK293T Cells were transfected with CRISPRa. Subsequently, qRT-PCR was used to assess the expression of *EYA1*, and normalized to the expressions of the *GAPDH* housekeeping gene. Data are presented as the mean ± SEM (n = 3). (*right*) Graphical depiction of *EYA1* gene’s promoter region including transcription start sites (TSS). gRNA1–gRNA4 represent target sites of CRISPRa. (c) Schematic representation of monoallelic and biallelic *EYA1* KO cells with their genotypes. *EYA1* haploinsufficiency was rescued using CRISPRa. (d) As in (b), except that dSaCas9- VP64 and either gRNA 2 or 3 were expressed in monoallelic and biallelic *EYA1* knockout HEK293T cells, as well as wild-type HEK293T cells. Values represent relative mRNA levels compared to HEK293 cells without transfection (control). (e) The *MYOG* promoter-driven luciferase reporter and the *SIX1* wild-type construct were introduced into HEK293T cells and variants with either monoallelic or biallelic *EYA1* knockouts. Relative luciferase activity (relative to the HEK293T control) is plotted as mean ± SEM (n = 3), *, *p* < 0.05, **, *p* < 0.01 (one-way ANOVA followed by Bonferroni’s multiple comparison test). (f) As in (d), except that whole cell extracts were subjected to SDS-polyacrylamide gel electrophoresis (PAGE) followed by immunoblotting (*Upper*). For quantification (*Lower*), signals from EYA1 were normalized to those of beta-actin. Bars represent the mean ± SEM. ***, *p* < 0.001 (n=3, two-tailed Student’s *t*-test).

In the ClinVar database (https://www.ncbi.nlm.nih.gov/clinvar/), we noted that 81 disease-causing *EYA1* variants (pathogenic or likely pathogenic) were implicated in BOR/BO syndrome (**Figure 5a and Supplementary Table 5**). The majority (68%) of the documented *EYA1* variants with BOR/BO phenotypes were loss-of-function, including nonsense, frameshift, canonical splicing, and SVs, resulting in haploinsufficiency (**Figure 5b**). Our results indicate that CRISPRa holds potential as a versatile genome editing approach to address *EYA1* haploinsufficiency, including those involving CGRs, suggesting its significance for personalized genome-specific interventions.

**Figure 5.**
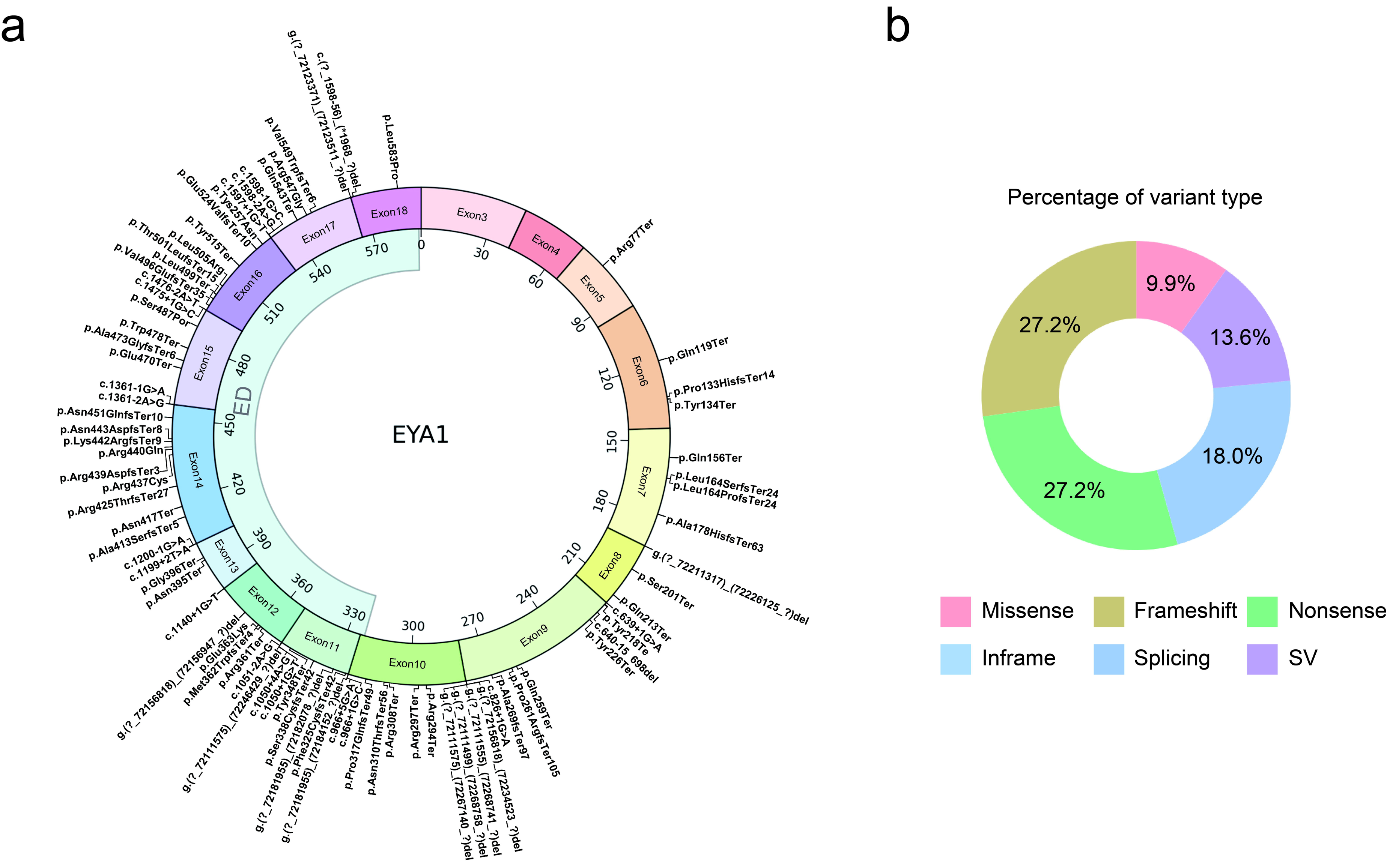
Genomic landscape of disease-causing *EYA1* variants implicated in BOR/BO syndrome. (a) Circos plot (https://github.com/SNUH-hEARgeneLab/WGS_analysis.) illustrating the distribution of all reported disease-causing *EYA1* variants from the ClinVar database. On the outer domain, the *EYA1* exons and corresponding reported variants are depicted, while the inner domain displays the EYA domain (ED) of EYA1. (b) Pie chart demonstrating the percentage of each variant type of *EYA1* disease- causing variants associated with BOR/BO syndrome.

## Discussion

This study highlights the importance of SVs, including CGRs and inversions, in rare diseases^1,26^. In the literature, various genomic rearrangements, such as cryptic inversions and large deletions, have been well-documented in approximately 20% of BOR/BO cases^24,27^. In certain cases, not only *EYA1* but also other genes were found to be involved, resulting in BOR/BO syndrome with additional clinical phenotypes^27^. Furthermore, non-allelic homologous recombination and human endogenous retrovirus elements are known to induce recurrent genomic rearrangements associated with BOR/BO syndrome^28,29^. The advancement of genomic technologies, coupled with reduced sequencing costs and improved data management, has made extraordinary strides toward deciphering the complex genetic architecture underlying human genetic disorders^30,31^. In parallel with this, precision genome editing approaches tailored to the patient’s genome facilitate the recovery of transcriptional activity vital for target gene expression. Therefore, we believe that the dual CRISPR/Cas9 system or CRISPRa-based gene therapy could have the potential to expand the treatment landscape for human genetic disorders, even for pathogenic SVs and CGRs.

Previous studies have shown the potential of targeted gene addition strategies as a therapeutic approach for Hemophilia A, which is often caused by pathogenic inversions of the *F8* gene^32^. This approach led to the restoration of *F8* expression in mesenchymal stem cells and endothelial cells, which had been differentiated from gene-corrected induced pluripotent stem cells (iPSCs)^32^. Moreover, Hu et al. provided a gene correction strategy for Hemophilia A, caused by an *F8* intron 1 large sequence inversion variant, through homology-mediated end joining with a high efficiency of 10.2%^33^. Genome editing using paired gRNAs has demonstrated efficient genomic modifications across various preclinical studies and even extended to human trials in the treatment of ophthalmic diseases^34^. The human trial used paired gRNAs to target and delete the aberrant splice donor site caused by the *CEP290* variant, which is commonly associated with Leber congenital amaurosis type 10 (LCA10), suggesting the potential of a Cas9 nuclease and paired gRNA system for therapeutic genome editing in human genetic disorders^34^. Furthermore, prime editing approaches that use paired pegRNAs (TwinPE)^35^ and prime editor nuclease-mediated translocation and inversion (PETI)^7^ can facilitate programmable genome editing in mammalian cells. These approaches utilize paired pegRNAs oriented in a protospacer adjacent motif (PAM)-in configuration to structure 3′ flaps on opposing genetic strands. These strategies demonstrate variances in both the synthesized flaps and the approaches used for DNA target incision^35^. Although CRISPR-based genome editing technologies to correct inversion sequences are evolving, there is a lack of published evidence on the therapeutic potential of Cas9 nuclease with paired gRNAs to target disease-causing paracentric inversions in patient-derived cells.

Cas9-derived DNA DSBs at dual loci within the *EYA1* gene have the potential to catalyze genomic rearrangements, as demonstrated by the paracentric inversion editing event in this study. Mechanistically, the CRISPR-Cas9 system, when used with paired gRNAs designed to target distinct genomic locations, has the potential to yield diverse modifications in DNA^36,37^. The inversion generated by a dual CRISPR/Cas9 system has been demonstrated across mammalian cell lines^38,39^ as well as *in vivo* in animal models^40^, suggesting the validity of the system in engineering specific inversion mutations. Encouraged by these insights, we hypothesize that paired gRNAs could be a potential therapeutic option to correct the disease-causing CGRs implicated in BOR/BO syndrome. We successfully applied the dual CRISPR/Cas9 system that specifically targeted a disease-causing paracentric inversion in patient-derived cells. The system demonstrated significant editing efficiency, leading to the restoration of significant gene expression and associated transcriptional functionality. Considering that an editing efficiency of even 1-2% in primary cells using CRISPR/Cas9 nuclease has been shown to rescue the hearing function of mice with *Atp2b2* mutations^41^, the 1.6% editing efficiency we achieved in patient-derived fibroblasts is particularly noteworthy. This result raises the possibility of restoring disease phenotypes, such as hearing impairment, in BOR/BO patients with *EYA1* CGRs including paracentric inversion.

Understanding the mechanistic pathways has the potential to significantly expand the therapeutic applications of the dual CRISPR/Cas9 system in clinical settings. When the Cas9 nuclease does not simultaneously cleave at both target sites, it instigates a sequence of events that leads to the accumulation of mutations around the gRNA target regions, which is attributed to the cellular repair mechanisms NHEJ and microhomology-mediated end joining ^42^. Alternatively, when Cas9 nucleases simultaneously cleave at both target loci the subsequent ligation could induce a large deletion, inversion, or translocation of the genomic fragment intervening between the target sites. Documented instances of such inversions suggest a scenario where both sgRNAs interact with the Cas protein, forming a single complex capable of excising gene segments^43^. This process facilitates the complex in generating a nick in both sgRNAs, thereby presenting three potential pathways for DNA repair. These include small indels, large deletion, substantial deletions, or inversions^44^. More specifically, it is plausible to posit that the formation of an inversion concurrently negates PAM sites and excises a fraction of the sgRNAs, liberating the cleavage sites^43^.

CRISPRa utilizes a Cas9 variant, dCas9, which lacks nuclease activity, combined with a transcriptional activator. By targeting the *EYA1* promoter, the dCas9-VP64 fusion used in our CRISPRa system increases *EYA1* expression (see **Figure 4b**), leading to significantly improved transcriptional activity essential for target gene expression. Of the documented *EYA1* variants linked with BOR/BO phenotypes, approximately 70% are described as monoallelic loss-of-function variants, including nonsense, frameshift, canonical splicing, and SVs (see **Figure 5b**). As a consequence, there is a state of haploinsufficiency. This suggests that CRISPRa-based gene therapies may offer substantial translational potential for approximately 70% of disease-causing *EYA1* variants caused by haploinsufficiency.

In conclusion, our results pave the way for the potential development of gene editing therapeutics for the clinical application of human genetic disorders caused by pathogenic SVs such as inversion and genomic rearrangements. In particular, BOR/BO syndrome stands as a representative case where hearing impairment is the most penetrant symptom^17,45^. Most branchial anomalies associated with this syndrome can be managed surgically, and the renal phenotype is rare. The inner ear presents a particularly promising target for gene therapy due to its low immunogenicity and compact enclosed structure that facilitates localized intervention. Previous studies have highlighted the efficacy of adeno-associated virus (AAVs) in transducing overexpressed cDNA and gene editing materials into target cells within the inner ear^46,47^. The advancements in gene editing technologies, including dual CRISPR/Cas9 system and CRISPRa, could expand the therapeutic landscape related to human genetic disorders, and encompass conditions such as BOR/BO syndrome due to *EYA1* CGRs associated with haploinsufficiency.

## Data Availability

All data produced in the present study are available upon reasonable request to the authors

## Acknowledgments

This research was supported and funded by SNUH Kun-hee Lee Child Cancer & Rare Disease Project, Republic of Korea (FP-2022-00001-004 to S-Y. Lee), National Research Foundation of Korea (NRF) and funded by the Ministry of Education (grant number: 2022R1C1C1003147 to Sang-Yeon Lee), SNUH Research Fund (04-2022- 4010 to S-Y. Lee and 04-2022-3070 to S-Y. Lee), National Research Foundation of Korea (2020R1A2C2101714 to D.K), Korean Fund for Regenerative Medicine (KFRM) grant funded by the Korean government (Ministry of Science and ICT, Ministry of Health & Welfare [21A0202L1-12 to D.K.]), and Korea Health Technology R&D Project through the Korea Health Industry Development Institute (KHIDI), funded by the Ministry of Health & Welfare, Republic of Korea (HI21C1314 and HR22C1363 to D.K.).

## Notes

### Competing Interest Statement

The authors have declared no competing interest.

### Author Declarations

Institutional Review Board of Seoul National University Hospital gaved ethical approval for this work (IRB-H-0905-041-281 and IRB-H-2202-045-1298).

