## Supplementary Materials for "CRISPR-based editing strategies to rectify *EYA1* complex genomic rearrangement linked to haploinsufficiency"

This PDF file includes:

Supplementary Figures 1-3

Supplementary Tables 1-5

**Supplementary Figure 1. Normal *EYA1* copy number state identified through MLPA.**

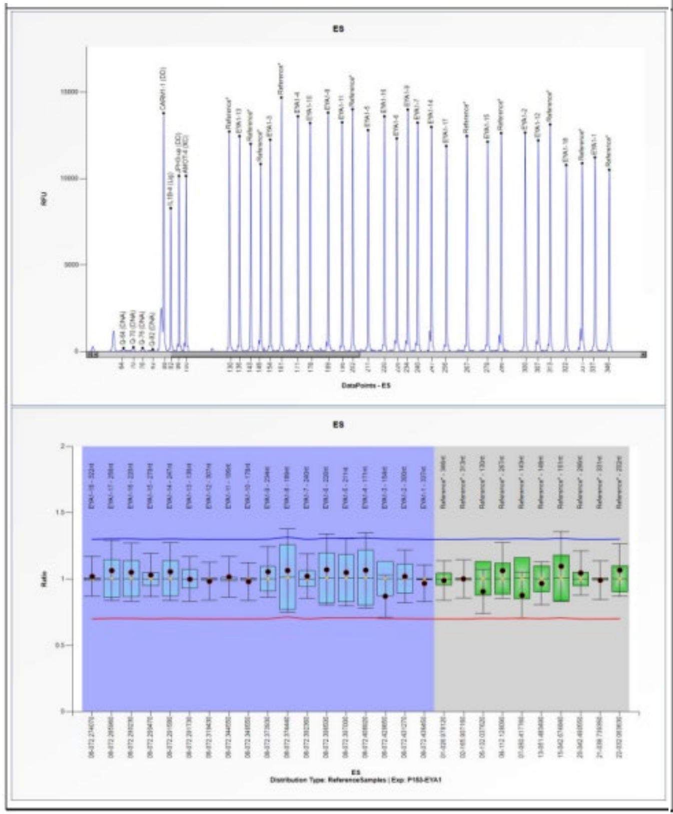


| D [nt] | Gene-Exon | Chr.band | hg18 loc. | Height | Area | Ratio[H] | Stdev | [REF] | Width | d[nt] | [Mut details] |
|---|---|---|---|---|---|---|---|---|---|---|---|
| 322 | EYA1-18 | 08q13.3 | 08-072.274070 | 10785 | 96529 | *1.02* | 0.08 | = | 59 | 0.0 | - |
| 256 | EYA1-17 | 08q13.3 | 08-072.285960 | 11885 | 96619 | *1.06* | **0.11** | = | 55 | 0.0 | - |
| 220 | EYA1-16 | 08q13.3 | 08-072.290230 | 13600 | 104863 | *1.05* | **0.11** | = | 52 | 0.1 | - |
| 279 | EYA1-15 | 08q13.3 | 08-072.290470 | 12134 | 102378 | *1.03* | 0.08 | = | 60 | 0.1 | - |
| 247 | EYA1-14 | 08q13.3 | 08-072.291580 | 12985 | **100330** | *1.06* | **0.11** | = | 29 | 0.0 | - |
| 136 | EYA1-13 | 08q13.3 | 08-072.291730 | 12457 | 91836 | *1* | 0.08 | = | 68 | 0.0 | - |
| 307 | EYA1-12 | 08q13.3 | 08-072.319430 | 12210 | 107088 | *0.98* | 0.07 | = | 69 | 0.1 | - |
| 196 | EYA1-11 | 08q13.3 | 08-072.344550 | 13254 | **95780** | *1.02* | 0.08 | = | 30 | 0.1 | - |
| 178 | EYA1-10 | 08q13.3 | 08-072.346550 | 13211 | 97845 | *0.98* | 0.07 | = | 71 | 0.0 | - |
| 234 | EYA1-9 | 08q13.3 | 08-072.373930 | 13998 | 114731 | *1.06* | 0.1 | = | 66 | 0.0 | - |
| 189 | EYA1-8 | 08q13.3 | 08-072.374440 | 13820 | 105034 | *1.07* | **0.16** | = | 51 | 0.0 | - |
| 240 | EYA1-7 | 08q13.3 | 08-072.392360 | 13232 | 106704 | *1.02* | 0.08 | = | 57 | 0.0 | - |
| 226 | EYA1-6 | 08q13.3 | 08-072.396500 | 12327 | **94592** | *1.07* | **0.13** | = | 37 | 0.0 | - |
| 211 | EYA1-5 | 08q13.3 | 08-072.397000 | 12804 | 97041 | *1.05* | **0.13** | = | 43 | 0.0 | - |
| 171 | EYA1-4 | 08q13.3 | 08-072.408920 | 13605 | 100138 | *1.07* | **0.14** | = | 45 | 0.0 | - |
| 154 | EYA1-3 | 08q13.3 | 08-072.429650 | 12252 | 91316 | *0.87* | 0.08 | = | 63 | 0.0 | - |
| 300 | EYA1-2 | 08q13.3 | 08-072.431270 | 12657 | 108309 | *1.02* | 0.1 | = | 55 | 0.1 | - |
| 337 | EYA1-1 | 08q13.3 | 08-072.436450 | 11227 | 104399 | *0.97* | 0.07 | = | 73 | 0.1 | - |
| 346 | Reference* | 01p36.11 | 01-026.978120 | 10515 | 99392 | *0.99* | 0.07 | = | 80 | 0.0 | - |
| 313 | Reference* | 02q24.3 | 02-165.907160 | 13136 | 115651 | *1* | 0.07 | = | 57 | 0.1 | - |
| 130 | Reference* | 05q31.1 | 05-132.037620 | 12724 | 92526 | *0.91* | 0.08 | = | 67 | 0.1 | - |
| 267 | Reference* | 06q21 | 06-112.128090 | 12463 | 104080 | *1.06* | 0.1 | = | 62 | 0.0 | - |
| 143 | Reference* | 07p12.2 | 07-050.417760 | 12011 | 86911 | *0.88* | 0.09 | = | 70 | 0.0 | - |
| 148 | Reference* | 13q14.3 | 13-051.483490 | 10847 | 84258 | *0.97* | 0.08 | = | 57 | 0.1 | - |
| 161 | Reference* | 15q15.3 | 15-042.674840 | 14695 | 105744 | *1.1* | **0.13** | = | 59 | 0.0 | - |
| 286 | Reference* | 20q13.12 | 20-042.490550 | 12634 | **103695** | *1.05* | 0.08 | = | 34 | 0.1 | - |
| 331 | Reference* | 21q22.2 | 21-038.739350 | 10891 | **94758** | *0.99* | 0.07 | = | 29 | 0.1 | - |
| 202 | Reference* | 22q12.3 | 22-032.063630 | 14016 | 105860 | *1.07* | 0.1 | = | 46 | 0.0 | - |
| **Median value all probe values:** | | | | **12646** | **100234** | **1.02** | **0.08** | | | **57** | **0.03** |

**Supplementary Figure 2. WGS-derived IGV snapshots of two novel SVs.**

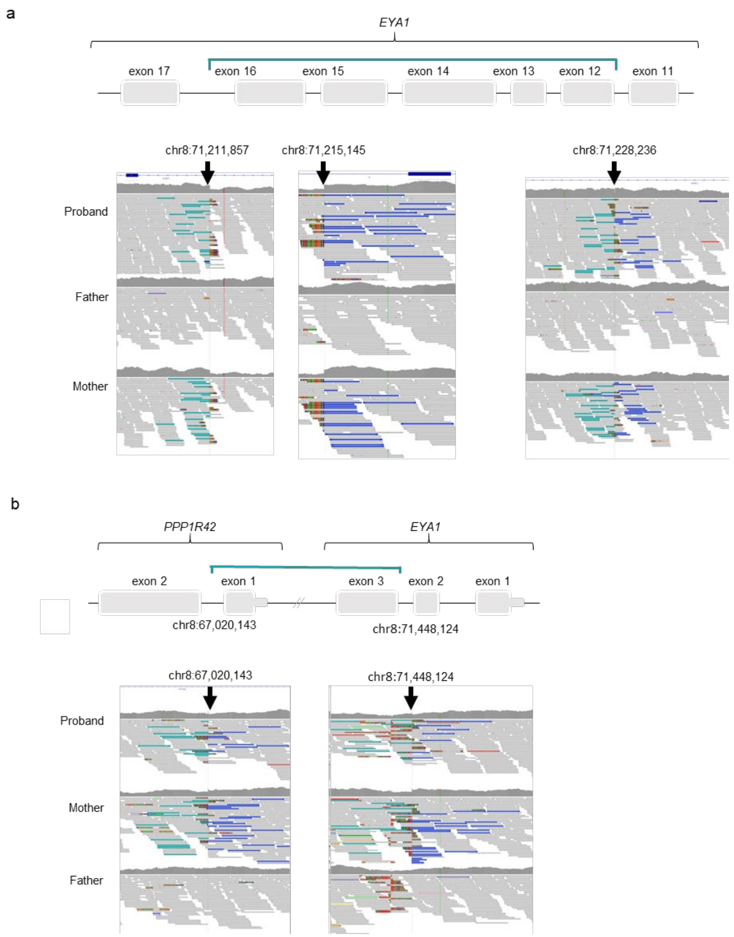


(a) CGRs (inversion with a deletion) where the junctions of three breakpoints were identified. The light green and blue lines show the joining of discordant reads for 3' to 3' and 5' to 5', respectively. (b) Cryptic large inversion where the junctions of two breakpoints were identified. The soft-clipped reads (light green and blue lines) exhibit a read pair overlaps with the inverted region.

**Supplementary Figure 3. Identification of two neo-junctional reads with microhomology.**

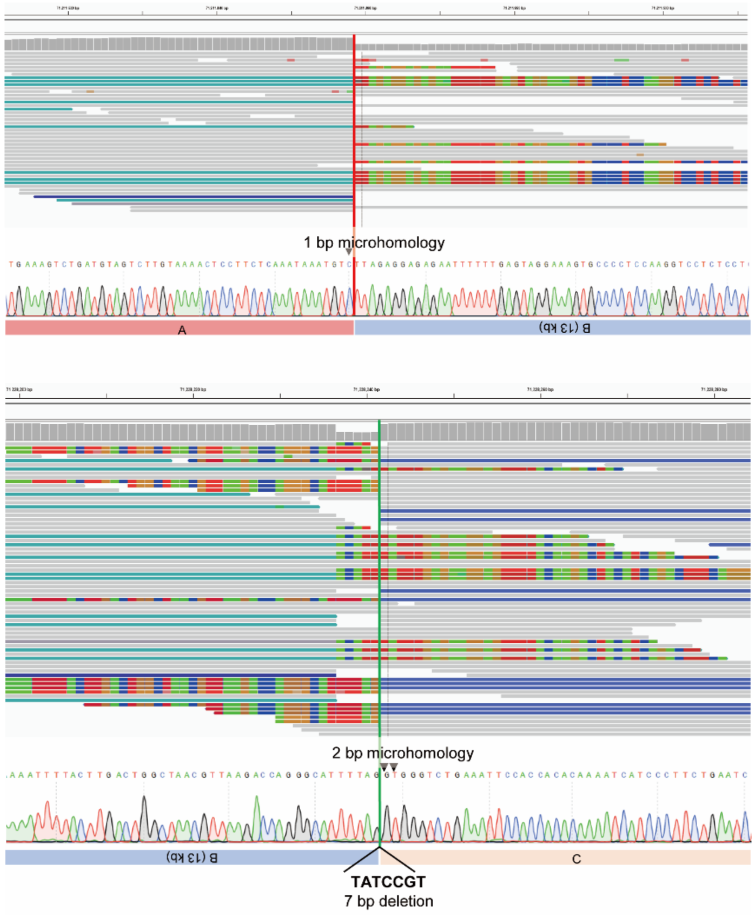


Two neo-junctional reads were validated with Sanger sequencing: one exhibiting 1 base pair (bp) of microhomology (junction A-B, position g.71215145) and the other showing 2 bp of microhomology (junction B-C, position g.71228236).

**Supplementary Table 1. List of primers employed to verify the locations of breakpoint junctions**

| Sample | Target | Primer Name | Primer sequence (5'-3') |
|---|---|---|---|
| EYA1 wild-type | A-B | EYA1_F1 | GTT CTA GCA AGA AAA CGC A |
| EYA1 wild-type | A-B | EYA1_R1 | ACA AGA ATA ATT TAA CCA ATT TGC |
| EYA1 wild-type | B-C | EYA1_F2 | TAG AGT TGT TTC ATT TCT AC |
| EYA1 wild-type | B-C | EYA1_R2 | ACT GGA TTT AGA CGT AAG AG |
| EYA1 complex SV | A-B inv | MUT-AB-F | CCA CTT GGG TTG TAT GTG CC |
| EYA1 complex SV | A-B inv | MUT-AB-R | GCC ATT ACA TGT CTA TAG ATT CA |
| EYA1 complex SV | B inv-C | MUT-BC-F | ACC AAT TTG CAA AAC TGG A |
| EYA1 complex SV | B inv-C | MUT-BC-R | CAT AGT GCA GTT CAC AC |

**Supplementary Table 2. List of primers employed to verify the locations of breakpoint junctions**

| Sample | Target | Primer Name | Primer sequence (5'-3') |
|---|---|---|---|
| SNUH | A-B | 67,020,143_WT_F | TCAGAACACATGGTGTGAAAACCT |
| SNUH | A-B | 67,020,143_WT_R | TGGATTATGTTGCCTCTTTGTGG |
| SNUH | B-C | 71,448,124_WT_F | TCAGGATGGAGGTTGCTGAA |
| SNUH | B-C | 71,448,124_WT_R | GCGAGAAGTCCTATTGTGCAT |
| SNUH | A-B inv | 67,020,143_Mut_F | GTCTCTTGGCTCCTTTTCCCA |
| SNUH | A-B inv | 67,020,143_Mut_R | TGAAGGTTGGGATGGCTGTG |
| SNUH | B inv-C | 71,448,124_Mut_F | TGGATTATGTTGCCTCTTTGTGG |
| SNUH | B inv-C | 71,448,124_Mut_R | TGCGAGAAGTCCTATTGTGC |

**Supplementary Table 3. List of primers employed to verify the locations of breakpoint junctions**

| Sample | Target | Primer Name | Primer sequence (5'-3') |
|---|---|---|---|
| WT | A-B | EYA1_C_F | ACGCCAACAGATATGGGAGG |
| Mut | | | |
| T1/T2 edited | | EYA1_B_R | TGTAGCGGAAGGCCAACTTT |
| T1/T3 edited | | | |

**Supplementary Table 4. In-house database of BOR/BO syndrome caused by *EYA1* variants**

| Sex/Age | Gene | Variant | Branchial anomalies | Preauricular pits | Hearing loss (Type) | Renal anomalies | External | Middle | Inner | EVA | Typical vs. Atypical[a] |
|---|---|---|---|---|---|---|---|---|---|---|---|
| F/16-20 | *EYA1* | c.1319G>A;p.Arg440Gln | | | | | | | | | Typical |
| M/16-20 | *EYA1* | c.1220G>A:p.Arg407Gln | | | | | | | | | Typical |
| F/6-10 | *EYA1* | c.1081C>T:p.Arg361Ter | | | | | | | | | Typical |
| M/6-10 | *EYA1* | c.1081C>T:p.Arg361Ter | | | | | | | | | Typical |
| M/6-10 | *EYA1* | c.1276G>A:p.Gly426Ser | | | | | | | | | Atypical |
| M/6-10 | *EYA1* | c.1276G>A:p.Gly426Ser | | | | | | | | | Atypical |
| F/11-15 | *EYA1* | Deletion | | | | | | | | | Typical |
| M/6-10 | *EYA1* | c.1081C>T:p.Arg361Ter | | | | | | | | | Typical |
| F/21-25 | *EYA1* | Inversion reciprocal with deletion | | | | | | | | | Typical |
| F/51-55 | *EYA1* | Inversion reciprocal with deletion | | | | | | | | | Atypical |
| F/26-30 | *EYA1* | Inversion reciprocal with deletion | | | | | | | | | Typical |
| M/31-35 | *EYA1* | Inversion reciprocal with deletion | | | | | | | | | Typical |
| M/31-35 | *EYA1* | c.1623_1626dup:p.Gln543AsnfsTer90 | | | | | | | | | Typical |
| M/21-25 | *EYA1* | c.1360G>T:p.Gly454Cys | | | | | | | | | Typical |
| F/6-10 | *EYA1* | c.1117_1118delCA:p.His373PhefsTer4 | | | | | | | | | Typical |
| F/0-5 | *EYA1* | EYA1 large genomic inversion | | | | | | | | | Typical |
| M/41-45 | *EYA1* | EYA1 large genomic inversion | | | | | | | | | Typical |

*EYA1*, canonical transcript NM_000503.6

Abbreviations: F, female; M, male; SNHL, sensorineural hearing loss; MHL, mixed hearing loss; EVA, enlarged vestibular aqueducts; N/A, not available.

[a]Note, if the note doesn't satisfy the standard criteria for BOR/BO syndrome (at least three major criteria or two major and two minor criteria), it can be classified as atypical BOR/BO syndrome

**Supplementary Table 5. Disease-causing *EYA1* variants associated with BOR/BO syndrome (pathogenic or likely pathogenic in ClinVar database)**

| No | Genomic Position (GRCh37/hg19) | HGVS | | Location (Exon/Domain) | ClinVar | Molecular Consequence |
|---|---|---|---|---|---|---|
| | | Nucleotide | Protein | | | |
| **1** | Chr8:72234477-G-A | c.229C>T | p.Arg77Ter | Exon 5 / Region:Disordered (1 - 95) | Pathogenic (PMID18220287) | Nonsense |
| **2** | Chr8:72234032-G-A | c.355C>T | p.Gln119Ter | Exon 6 / Absent | Pathogenic (PMID18220287) | Nonsense |
| **3** | Chr8:72233981-TGCCGTACG-T | c.398_405del | p.Pro133HisfsTer14 | Exon 6 / Absent | Pathogenic (PMID9359046) | Frameshift |
| **4** | Chr8:72233985-G-C | c.402C>G | p.Tyr134Ter | Exon 6 / Absent | Pathogenic (PMID21280147) | Nonsense |
| **5** | Chr8:72229877-G-A | c.466C>T | p.Gln156Ter | Exon 7 / Absent | Pathogenic (PMID22447252) | Nonsense |
| **6** | Chr8:72229853-G-GA | c.489dup | p.Leu164SerfsTer24 | Exon 7 / Absent | Pathogenic (PMID10991693) | Frameshift |
| **7** | Chr8:72229852-A-AG | c.490dup | p.Leu164ProfsTer24 | Exon 7 / Absent | Pathogenic (PMID18220287) | Frameshift |
| **8** | Chr8:7229810-GC-G | c.532del | p.Ala178HisfsTer63 | Exon 7 / Absent | Pathogenic (PMID18220287) | Frameshift |
| **9** | Chr8:72211317-72226125 | g.(?_72211317)_(72226125_?)del | g.(?_72211317)_(72226125_?)del | Intron 7 | Pathogenic (PMID21280147) | SV |
| **10** | Chr8:72211910-G-C | c.602C>G | p.Ser201Ter | Exon 8 / Absent | Pathogenic (PMID18220287) | Nonsense |
| **11** | Chr8:72211875-G-A | c.637C>T | p.Gln213Ter | Exon 8 / Absent | Pathogenic (PMID18220287) | Nonsense |
| **12** | Chr8:72211872-C-T | c.639+1G>A | p.? | Intron 8 | Pathogenic (PMID18220287) | Splicing |
| **13** | Chr8:72211454-A-C | c.654T>G | p.Tyr218Ter | Exon 9 / Absent | Pathogenic (PMID19206155) | Nonsense |

| | | | | | | |
|---|---|---|---|---|---|---|
| **14** | Chr8:72211410-72211483 | c.640-15_698del | p.? | Exon 9 / Absent | Pathogenic (PMID18220287) | Splicing |
| **15** | Chr8:72211430-G-C | c.678C>G | p.Tyr226Ter | Exon 9 / Absent | Pathogenic (PMID18220287) | Nonsense |
| **16** | Chr8:72211333-G-A | c.775C>T | p.Gln259Ter | Exon 9 / Region:Disordered (240 - 320) | Pathogenic (PMID18220287) | Nonsense |
| **17** | Chr8:72211325CG-C | c.782del | p.Pro261ArgfsTer105 | Exon 9 / Region:Disordered (240 - 320) | Pathogenic (PMID18220287) | Frameshift |
| **18** | Chr8:72211301-TG-T | c.806del | p.Ala269fsTer97 | Exon 9 / Region:Disordered (240 - 320) | Pathogenic (PMID18220287) | Frameshift |
| **19** | Chr8:72211281-C-T | c.826+1G>A | p.? | Intron 9 | Likely Pathogenic (PMID18220287) | Splicing |
| **20** | Chr8:72156818-72234523 | g.(?_72156818)_(72234523_?)del | g.(?_72156818)_(72234523_?)del | Intron 9 | Pathogenic (PMID18220287) | SV |
| **21** | Chr8:72111555-72268741 | g.(?_72111555)_(72268741_?)del | g.(?_72111555)_(72268741_?)del | Intron 9 | Pathogenic (PMID18220287) | SV |
| **22** | Chr8:72111499-72268758 | g.(?_72111499)_(72268758_?)del | g.(?_72111499)_(72268758_?)del | Intron 9 | Pathogenic (PMID25135225) | SV |
| **23** | Chr8:72111575-72267140 | g.(?_72111575)_(72267140_?)del | g.(?_72111575)_(72267140_?)del | Intron 9 | Pathogenic (PMID31427586) | SV |
| **24** | Chr8:72184079-G-A | c.880C>T | p.Arg294Ter | Exon 10 / Region:Disordered (240 - 320) | Pathogenic (PMID35982127) | Nonsense |
| **25** | Chr8:72184070-G-A | c.889C>T | p.Arg297Ter | Exon 10 / Region:Disordered (240 - 320) | Pathogenic (PMID18220287) | Nonsense |
| **26** | Chr8:72184037-G-A | c.922C>T | p.Arg308Ter | Exon 10 / Region:Disordered (240 - 320) | Pathogenic (PMID29500469) | Nonsense |
| **27** | Chr8:72184029-GT-G | c.929del | p.Asn310ThrfsTer56 | Exon 10 / Region:Disordered (240 - 320) | Pathogenic (PMID33532864) | Frameshift |

| | | | | | | |
|---|---|---|---|---|---|---|
| **28** | Chr8:72184009-TG-T | c.950del | p.Pro317GlnfsTer49 | Exon 10 / Region:Disordered (240 - 320) | Pathogenic (PMID18220287) | Frameshift |
| **29** | Chr8:72183992-C-G | c.966+1G>C | p.? | Intron 10 | Likely Pathogenic (PMID30655312) | Splicing |
| **30** | Chr8:72183988-C-T | c.966+5G>A | p.? | Intron 10 | Pathogenic (PMID27657687) | Splicing |
| **31** | Chr8:72181955-72184152 | g.(?_72181955)_(72184152_?)del | g.(?_72181955)_(72184152_?)del | Intron 10 | Pathogenic (PMID18220287) | SV |
| **32** | Chr8:72182051-A-AAC | c.972_973dup | p.Phe325CysfsTer42 | Exon 11 / Absent | Pathogenic (PMID9359046) | Frameshift |
| **33** | Chr8:72181955-72182078 | g.(?_72181955)_(72182078_?)del | g.(?_72181955)_(72182078_?)del | Exon 11 / Absent | Pathogenic (PMID19206155) | SV |
| **34** | Chr8:72182008-CAAGG-C | c.1013_1016del | p.Ser338CysfsTer27 | Exon 11 / Absent | Pathogenic (PMID18220287) | Frameshift |
| **35** | Chr8:72181981-A-C | c.1044T>G | p.Tyr348Ter | Exon 11 / Absent | Pathogenic (PMID18220287) | Nonsense |
| **36** | Chr8:72181974-C-A | c.1050+1G>T | p.? | Intron 11 | Pathogenic (PMID18220287) | Splicing |
| **37** | Chr8:72181971-T-C | c.1050+4A>G | p.? | Intron 11 | Pathogenic (PMID17576681) | Splicing |
| **38** | Chr8:72111575-72246429 | g.(?_72111575)_(72246429_?)del | g.(?_72111575)_(72246429_?)del | Intron 11 | Pathogenic (PMID28832562) | SV |
| **39** | Chr8:72156929-T-C | c.1051-2A>G | p.? | Intron 11 | Pathogenic (PMID21280147) | Splicing |
| **40** | Chr8:72156897-G-A | c.1081C>T | p.Arg361Ter | Exon 12 / Absent | Pathogenic (PMID26969326) | Nonsense |
| **41** | Chr8:72156893-AT-A | c.1084del | p.Met362TrpfsTer4 | Exon 12 / Absent | Pathogenic (PMID18220287) | Frameshift |
| **42** | Chr8:72156891-C-T | c.1087G>A | p.Glu363Lys | Exon 12 / Absent | Pathogenic (PMID10655545) | Missense |

| | | | | | | |
|---|---|---|---|---|---|---|
| **43** | Chr8:72156818-72156947 | g.(?_72156818)_(72156947_?)del | g.(?_72156818)_(72156947_?)del | Exon 12 / Absent | Pathogenic (PMID30937553) | SV |
| **44** | Chr8:72156837-C-A | c.1140+1G>T | p.? | Intron 12 | Pathogenic (PMID23840632) | Splicing |
| **45** | Chr8:72129216-T-TA | c.1182dup | p.Asn395Ter | Exon 13 / Absent | Pathogenic (PMID18220287) | Nonsense |
| **46** | Chr8:72129213-C-A | c.1186G>T | p.Gly396Ter | Exon 13 / Absent | Pathogenic (PMID18220287) | Nonsense |
| **47** | Chr8:72129198-A-T | c.1199+2T>A | p.? | Intron 13 | Likely Pathogenic (PMID18220287) | Splicing |
| **48** | Chr8:72129088-C-T | c.1200-1G>A | p.? | Intron 13 | Pathogenic (PMID18220287) | Splicing |
| **49** | Chr8:72129050-C-CT | c.1236dup | p.Ala413SerfsTer5 | Exon 14 / Absent | Pathogenic (PMID18220287) | Frameshift |
| **50** | Chr8:72129038-T-TA | c.1248dup | p.Asn417Ter | Exon 14 / Absent | Pathogenic (PMID18220287) | Nonsense |
| **51** | Chr8:72129014-G-GT | c.1272dup | p.Arg425ThrfsTer27 | Exon 14 / Absent | Pathogenic (PMID18220287) | Frameshift |
| **52** | Chr8:72128978-G-A | c.1309C>T | p.Arg437Cys | Exon 14 / Absent | Likely Pathogenic (PMID34031707) | Missense |
| **53** | Chr8:72128972-CT-C | c.1315del | p.Arg439AspfsTer3 | Exon 14 / Absent | Pathogenic (PMID18220287) | Frameshift |
| **54** | Chr8:72128968-C-T | c.1319G>A | p.Arg440Gln | Exon 14 / Absent | Pathogenic (PMID30655312) | Missense |
| **55** | Chr8:72128960-CTT-C | c.1325_1326del | p.Lys442ArgfsTer9 | Exon 14 / Absent | Pathogenic (PMID19206155) | Frameshift |
| **56** | Chr8:72128956-ATC-A | c.1329_1330del | p.Glu443AspfsTer8 | Exon 14 / Absent | Pathogenic (PMID18220287) | Frameshift |
| **57** | Chr8:72128937-A-GG | c.1350delinsCC | p.Asn451GlnfsTer10 | Exon 14 / Absent | Pathogenic (PMID9020840) | Frameshift |

| | | | | | | |
|---|---|---|---|---|---|---|
| **58** | Chr8:72127965-T-C | c.1361-2A>G | p.? | Intron 14 | Pathogenic (PMID18220287) | Splicing |
| **59** | Chr8:72127964-C-T | c.1361-1G>A | p.? | Intron 14 | Likely Pathogenic (PMID18220287) | Splicing |
| **60** | Chr8:72127916-C-A | c.1408G>T | p.Glu470Ter | Exon 15 / Absent | Pathogenic (PMID25741868) | Nonsense |
| **61** | Chr8:72127902-CAGGG-C | c.1418_1421del | p.Ala473GlyfsTer6 | Exon 15 / Absent | Pathogenic (PMID18220287) | Frameshift |
| **62** | Chr8:72127891-C-T | c.1433G>A | p.Trp478Ter | Exon 15 / Absent | Pathogenic (PMID18220287) | Nonsense |
| **63** | Chr8:72127865-A-G | c.1459T>C | p.Ser487Pro | Exon 15 / Absent | Pathogenic (PMID9361030) | Missense |
| **64** | Chr8:72127848-C-G | c.1475+1G>C | p.? | Intron 15 | Pathogenic (PMID23506628) | Splicing |
| **65** | Chr8:72127745-T-A | c.1476-2A>T | p.? | Intron 15 | Pathogenic (PMID21280147) | Splicing |
| **66** | Chr8:72127730-TCA-T | c.1487_1488del | p.Val496GlufsTer35 | Exon 16 / Absent | Pathogenic (PMID18220287) | Frameshift |
| **67** | Chr8:72127719-TACTA-T | c.1496_1499del | p.Leu499Ter | Exon 16 / Absent | Pathogenic (PMID35005812) | Nonsense |
| **68** | Chr8:72127711-GTAGTTGT-G | c.1501_1507del | p.Thr501LeufsTer15 | Exon 16 / Absent | Pathogenic (PMID15493068) | Frameshift |
| **69** | Chr8:72127705-A-C | c.1514T>G | p.Leu505Arg | Exon 16 / Absent | Pathogenic (PMID9361030) | Missense |
| **70** | Chr8:72127674-A-T | c.1545T>A | p.Tyr515Ter | Exon 16 / Absent | Pathogenic (PMID18220287) | Nonsense |
| **71** | Chr8:72127648-T-TTA | c.1570_1571insTA | p.Glu524ValfsTer10 | Exon 16 / Absent | Pathogenic (PMID18220287) | Frameshift |
| **72** | Chr8:72127640-A-T | c.1579T>A | p.Tys527Asn | Exon 16 / Absent | Likely Pathogenic (PMID23435380) | Missense |

| | | | | | | |
|---|---|---|---|---|---|---|
| **73** | Chr8:72127621-C-A | c.1597+1G>T | p.? | Intron 16 | Likely Pathogenic (PMID18220287) | Splicing |
| **74** | Chr8:72123493-T-C | c.1598-2A>G | p.? | Intron 16 | Pathogenic (PMID23840632) | Splicing |
| **75** | Chr8:72123492-C-G | c.1598-1G>C | p.? | Intron 16 | Pathogenic (PMID18220287) | Splicing |
| **76** | Chr8:72123462-G-A | c.1627C>T | p.Gln543Ter | Exon17 / Absent | Pathogenic (PMID34160378) | Nonsense |
| **77** | Chr8:72123450-T-C | c.1639A>G | p.Arg547Gly | Exon17 / Absent | Pathogenic (PMID10655545) | Missense |
| **78** | Chr8:72123444-CT-C | c.1644del | p.Val549TrpfsTer6 | Exon17 / Absent | Pathogenic (PMID18220287) | Frameshift |
| **79** | Chr8:72123371-72123511 | g.(?_72123371)_(72123511_?)del | g.(?_72123371)_(72123511_?)del | Exon17 / Absent | Pathogenic (PMID29500469) | SV |
| **80** | Chr8:72109607-72123547 | c.(?_1598-56)_(*1968_?)del | c.(?_1598-56)_(*1968_?)del | Intron 17 | Pathogenic (PMID23851940) | SV |
| **81** | Chr8-72111606-A-G | c.1748T>C | p.Leu583Pro | Exon 18 / Absent | Likely Pathogenic (PMID31049720) | Missense |
